# Evidence of artemisinin partial resistance in North-western Tanzania: clinical and drug resistance markers study

**DOI:** 10.1101/2024.01.31.24301954

**Authors:** Deus S. Ishengoma, Celine I. Mandara, Catherine Bakari, Abebe A. Fola, Rashid A. Madebe, Misago D. Seth, Filbert Francis, Creyton Buguzi, Ramadhan Moshi, Issa Garimo, Samwel Lazaro, Abdallah Lusasi, Sijenunu Aaron, Frank Chacky, Ally Mohamed, Ritha J. A. Njau, Jovin Kitau, Charlotte Rasmussen, Jeffrey A. Bailey, Jonathan J. Juliano, Marian Warsame

**Affiliations:** National Institute for Medical Research, Dar es Salaam, Tanzania; Harvard T.H Chan School of Public Health, Boston, MA, USA; Faculty of Pharmaceutical Sciences, Monash University, VIC, Australia; Department of Biochemistry, Kampala International University in Tanzania, Dar es Salaam, Tanzania; Department of Pathology and Laboratory Medicine and Center for Computational Molecular Biology, Brown University, Providence, RI, USA; National Malaria Control Program (NMCP), Dodoma, Tanzania; Malariologist and Public Health Specialist, Muhimbili University of Health and Allied Sciences, School of Public Health and Social Sciences, Dar es Salaam, Tanzania; World Health Organization Country Office, Dar es Salaam, Tanzania; World Health Organization, Geneva, Switzerland; University of North Carolina, Chapel Hill, NC, USA; Gothenburg University, Gothenburg, Sweden; Benadir University, Mogadishu, Somalia

**Keywords:** Artemisinin partial resistance, artemisinin-based combination therapy, falcipar um malaria, Plasmodium falciparum, Kelch 13, Kagera, Tanzania

## Abstract

**Background:** Artemisinin-based combination therapies (ACTs) are the recommended antimalarial drugs for the treatment of uncomplicated malaria. The recent emergence of artemisinin partial resistance (ART-R) in Rwanda, Uganda and Eritrea is of great concern. In Tanzania, a nationwide molecular malaria surveillance in 2021 showed a high prevalence of the Kelch13 (K13) 561H mutation in Plasmodium falciparum from the north-western region, close to the border with Rwanda and Uganda. This study was conducted in 2022 to evaluate the efficacy of artemether-lumefantrine (AL) and artesunate-amodiaquine (ASAQ) for the treatment of uncomplicated falciparum malaria and to confirm the presence of ART-R in Tanzania.

**Methods:** This single-arm study evaluated the efficacy of AL and ASAQ in eligible children aged six months to 10 years at Bukangara Dispensary in Karagwe District, Kagera Region. Clinical and parasitological responses were monitored for 28 days according to standard WHO protocol. Mutations in K13 gene and extended haplotypes with these mutations were analysed using Sanger and whole genome sequencing data, respectively.

**Findings:** 176 children (88 in each AL and ASAQ group) were enrolled and all achieved the defined outcomes. PCR-corrected adequate clinical and parasitological response (ACPR) was 98.3% (95% CI: 90.8-100) and 100.0% (95% CI: 95.8-100) for AL and ASAQ, respectively. Parasitaemia on day 3 was observed in 11/88 (12.5%) and 17/88 (19.3%) in the AL and ASAQ groups, respectively. The half-life of parasitaemia was significantly higher (>6.5 hrs) in patients with parasitaemia on day 3 and/or mutations in K13 gene at enrolment. Most patients with parasitaemia on day 3 (8/11 = 72.7% in the AL group and 10/17 = 58.8% in the ASAQ group) had 561H mutation at enrolment. The parasites with K13 mutations were not similar to those from south-east Asia and Rwanda, but had the same core haplotype of a new 561H haplotype reported in Kagera in 2021.

**Interpretation:** These findings confirm the presence of ART-R in Tanzania. A context-specific strategy to respond to artemisinin partial resistance is urgently needed. Although both AL and ASAQ showed high efficacy, increased vigilance for reduced efficacy of these ACTs and detection of ART-R in other parts of the country is critical.

**Funding:** Bill and Melinda Gates Foundation to the World Health Organization (WHO, OPP 1209843) and the National Institute for Medical Research (NIMR, Inv. No. 002202), and US National Institute for Health (R01AI156267 to JAB, DSI and JJJ, and K24AI134990 to JJJ).

**Research in context:** *Evidence before this study:* Artemisinin partial resistance (ART-R) is defined as delayed clearance after treatment with an artemisinin combination therapy (ACT) or artesunate monotherapy of a parasite strain carrying a validated marker of ART-R. At present, 13 different Kelch13 (K13) mutations have been validated as markers of ART-R. ART-R is confirmed in an area if a quality-controlled study using an ACT or artesunate monotherapy, finds more than 5% of patients have parasites with validated K13 mutations and delayed clearance as evidenced by either persistent parasitemia detected by microscopy on day 3 or a parasite clearance half-life of ≥5 hours. ART-R was first reported from Cambodia in 2008 and later from several countries in Southeast Asia. Published articles up to December 2023 were searched in PubMed with the terms; “artemisinin”, “artemisinin partial resistance”, “artemisinin-based combination therapies”, “Kelch 13” in combination with “Africa” or “Tanzania”. The publications confirmed the emergence of ART-R associated with mutations in K13: 561H in Rwanda, A675V and C469Y in Uganda and R622I in Eritrea. All these studies showed a high cure rate of the tested ACTs. The R622I mutant was not reported from Southeast Asia but is circulating in the Horn of Africa (Eritrea, Ethiopia, Sudan and Somalia). In Tanzania, a nationwide malaria molecular surveillance launched in January 2021 showed a high prevalence of 561H mutation in the north-western region of Kagera, close to the border with Rwanda and Uganda.

*Added value of this study:* The study documented delayed parasite clearance associated with pre-treatment validated K13 561H mutation. It confirms and provides evidence for the first-time of ART-R in Kagera region, north-western Tanzania, an area close to the border with Rwanda and Uganda. This makes Tanzania the fourth country in Africa with confirmed ART-R. The study documents presence of K13 mutation associated with ART-R suggesting that partial resistance to artemisinins is rapidly evolving and can still be found in more areas of Africa. Parasites with K13 mutations were not similar to those from south-east Asia and Rwanda, but had the same core haplotype of a new 561H haplotype reported in Kagera in 2021.The findings of this study furthermore show that both AL and ASAQ are highly effective.

*Implications of all the available evidence:* The emergence of confirmed ART-R in Africa, so far in four countries (Rwanda, Uganda, Eritrea and Tanzania), poses a serious threat to malaria control in Africa, which accounts for more than 95% of the global malaria burden. The current evidence of ART-R in Kagera region calls for an urgent response, including the development of a context-specific strategy based on the recently launched WHO strategy to respond to antimalarial drug resistance in Africa. The fact that ART-R has been confirmed in Kagera region, an area bordering Rwanda and Uganda, where resistance also has been reported, also calls for cross-border collaboration to harmonize strategies to combat this threat in the Great Lakes region of Africa. Nationwide studies on molecular markers in Tanzania, which revealed a high prevalence of K13 validated mutations in the Kagera region, guided where to conduct the current study. This suggests that molecular marker surveillance could play an important role in conducting targeted antimalarial drug efficacy studies and confirming ART-R in other parts of Tanzania and beyond.

## Introduction

In response to the emergence and spread of *Plasmodium falciparum* resistance to chloroquine and then to sulfadoxine-pyrimethamine (SP), the World Health Organisation (WHO) recommended artemisinin-based combination therapies (ACTs) for the treatment of uncomplicated *P. falciparum* infections in 2001.^1^ Currently six ACTs recommended by WHO are artemether-lumefantrine (AL), artesunate-amodiaquine (ASAQ), dihydroartemisini n-piperaquine (DP), artesunate-mefloquine (ASMQ), artesunate-sulfadoxine-pyrimethamine (ASSP) and artesunate-pyronaridine (AP). The WHO recommends monitoring the efficacy of nationally recommended ACTs at least every two years at sentinel sites using the standard WHO protocol to inform treatment policy.^2^ High efficacy (over 90%) of AL has been reported in most studies in Africa.^3^ However, efficacy of AL below the 90% threshold recommended for treatment policy change was reported in Angola, Burkina Faso, the Democratic Republic of Congo and Uganda.^4–7^ It is worth noting, however, that these studies deviated from the WHO recommended PCR-correction upon which the 90% cut-off point was based.^8^

Artemisinin partial resistance (ART-R) is confirmed in an area when more than 5% of patients treated with an ACT or artesunate monotherapy harbour mutations validated to be associated with resistance and have delayed clearance described as either persistent parasitaemia at day 3 or a parasite clearance half-life of ≥5 hours.^9^ To date, 13 Kelch13 (K13) mutations have been validated to be associated with artemisinin partial resistance (F446I, N458Y, M476I, Y493H, R539T, I543T, P553L, R561H, P574L, C580Y, C469Y, R622I, A675V) and nine K13 mutations are considered candidate/associated markers (P441L, G449A, C469F, A481V, R515K, P527H, N537I/D, G538V, V568G).^3^ ART-R was first reported in Cambodia and later in several Southeast Asian countries.^10,11^ More recently, ART-R has been reported in Africa, in Rwanda, Uganda and Eritrea.^12–14^ Although none of the studies reported an associated reduced efficacy of the ACTs tested, the emergence of ART-R in Africa is of great concern and requires urgent attention. The WHO has recently launched a strategy to respond to antimalarial drug resistance in Africa, and countries are urged to develop a context-specific strategy to respond to this new threat.^9^

Tanzania was responsible for 3.2% and 4.4% of global malaria cases and deaths, respectively, in 2022 and is among the four countries responsible for more than 50% of malaria deaths globally, alongside Nigeria (31.1%), the Democratic Republic of Congo (11.6%) and Niger (5.6%).^15^ The country introduced AL as a first-line treatment in 2006.^16^ Artesunate-amodiaquine (ASAQ) is also recommended as an alternative ACT for the treatment of uncomplicated malaria if AL treatment fails or is contraindicated.^17^ Routine efficacy monitoring of AL and ASAQ has shown high efficacy to date.^18–20^ However, recent nationwide malaria molecular surveillance (MMS) studies conducted in 2021, covering 13 regions of the country, reported a hotspot with high prevalence of K13 561H mutations in Karagwe district in Kagera region.^21^ This finding prompted an immediate response through this study to assess the efficacy of AL and ASAQ treatments and confirm the presence of ART-R, in an area along the Kagera River and close to Rwanda and Uganda with significant cross-border population movements.^22^

## Methods

### Study design, site and population

This was a dual single-arm prospective study to evaluate the efficacy and safety of AL and ASAQ in children with uncomplicated falciparum infections. Children who met the study inclusion criteria were enrolled, treated with either AL or ASAQ under direct supervision and followed up for 28 days according to standard WHO protocol.^2^ The study was conducted from April to September 2022 at Bukangara dispensary in Karagwe District, Kagera Region. Karagwe district, with a population of 385,744 according to the 2022 census, borders Uganda to the north and Rwanda to the west, the districts of Misenyi and Muleba to the east, and Ngara and Biharamulo to the south (Fig. 1). The study site is located about 100 km from Rukara health facility in Kayonza district in Rwanda, where ART-R was confirmed in 2018.^12^ Malaria transmission in the study area usually peaks after the rainy season, between May and July and December and January.

**Figure 1:**
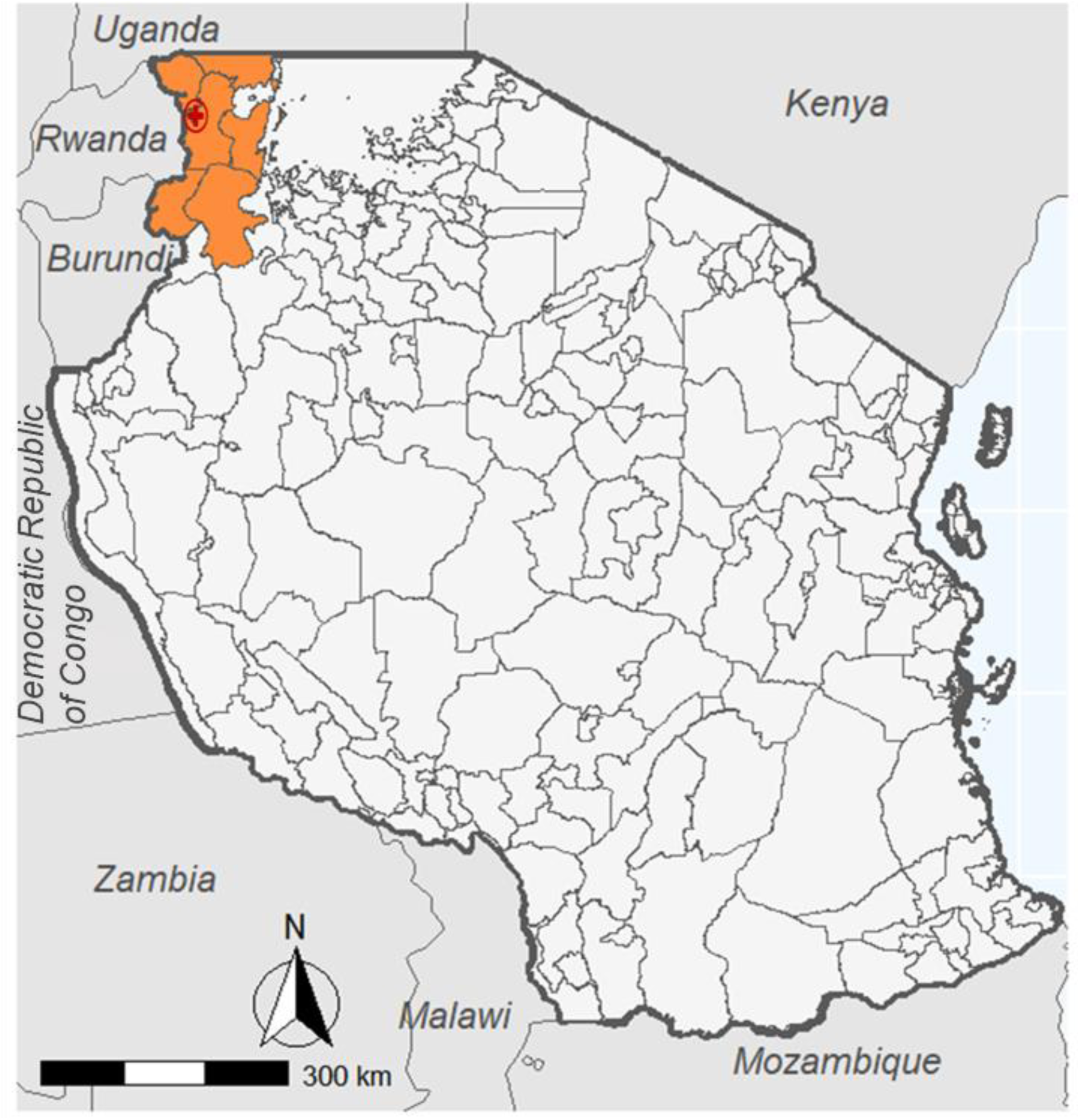
Map showing the location of Bukangara Dispensary in Karagwe district of Kagera region, North-western Tanzania.

### Procedures

#### Screening and recruitment

Children with suspected uncomplicated malaria were screened and enrolled if they had the following inclusion criteria: age between six months and 10 years, axillary temperature ≥ 37.5 °C) and/or reported history of fever in the last 24 hrs, confirmed *P. falciparum* mono-infec t ion with an asexual parasitaemia of 500-200,000/μl blood, haemoglobin levels ≥8 g/dL, and able and willing to attend scheduled follow-up visits. Children with the following exclusion criteria, based on WHO standard protocol^2^, were not recruited: general danger signs or symptoms of severe falciparum malaria as defined by WHO, mixed or mono infections with non-falciparum species, severe malnutrition (children 6-60 months), co-infections, anaemia (haemoglobin levels <8 g/dL), and history of drug reactions. Infected patients not eligible for enrolment in the study were treated according to the national treatment guideline.

#### Treatment and follow-up procedures

Children in the AL group received a dose twice daily for three consecutive days based on the recommended weight bands: one tablet for 5-14 kg body weight, two tablets for 15-24 kg body weight, three tablets for 25-34 kg body weight and four tablets for ≥35 kg body weight. Although all patients were provided with meals during their stay at the study health facility, no food was given at the time of drug administration. The ASAQ group received a daily dose for three days based on the following weight bands: one tablet of 25 artesunate/67.5 amodiaquine for 4.5 to >9 kg body weight, one tablet of 50 artesunate/135 amodiaquine for 9 −>18 kg body weight, one tablet of 100 artesunate/270 amodiaquine for 18 to <36 kg body weight. WHO prequalified AL (Cipla Ltd, India) and ASAQ (Winthrop®, Sanofi Aventis, Morocco) were provided by WHO headquarters. The study children were retained at the study facility for three days. All doses were administered orally and timely under direct observation and observed for vomiting for 30 minutes. If vomiting occurred within 30 minutes, another full dose was administered. Any patient who persistently vomited the study drug was withdrawn and treated with injectable artesunate followed by a full dose of AL according to the national treatment guidelines.^8^ Clinical and parasitological assessments were performed at scheduled visits (days 1, 2, 3, 7, 14, 21, and 28). In addition, parents/guardians were informed that they could bring their children back to the clinic at any time (unscheduled visits) if the child felt unwell. Children who did not come for their scheduled visits were visited at home to perform the scheduled assessments. Study drug safety was assessed at each follow-up visit by asking parents/guardians about the occurrence and nature of adverse events (AEs) and serious adverse events (SAEs), defined according to the WHO protocol 2009.^2^ Any observed serious adverse event was to be reported within 24 hours to the National Institute for Medical Research (NIMR), WHO, and the Medical Research Coordinating Committee of NIMR (NIMR-MRCC). Patients with AEs or SAEs were thoroughly investigated and managed.

#### Microscopy examination and Haemoglobin blood level

Thick and thin blood smears were taken through finger prick and Giemsa stained and examined to detect the presence of *P. falciparum* and to estimate parasite density before treatment (day 0) and at each scheduled (days 1, 3, 7, 14, 21 and 28) or unscheduled visit according to WHO protocol 2009.^2^ In addition, blood smears were taken every 8 hours until two consecutive smears were negative or the patient reached day 3 before clearing the parasites to assess parasite clearance time. All blood slides were read by two independent microscopists and the two parasite densities were averaged. If there were discrepancies in parasite positivity, parasite species or difference in parasite density of more than 50% between the two readings, the slide was re-examined by a third independent microscopist. A slide was declared negative if no parasites were seen after counting 1000 leukocytes. The presence of gametocytes at inclusion in the study or at follow-up visits was also recorded. In addition, 100 fields of the thick smears were examined on day 0 to rule out mixed infections. In cases of doubt, the thin film was examined for confirmation. The Hb level (g/dL) was measured with a HaemoCue® machine (HemoCue, Ångelholm, Sweden) to determine patients with anaemia (Hb <8g /dl) who were excluded from the study.

#### Parasite genotyping

Filter paper blood samples on Whatman #3 were collected as dried blood spot (DBS) from each patient on day 0 and on the day of parasite recurrence (from day 7 onwards). They were dried, stored in individual plastic bags with desiccant until analysis, protecting them from light, moisture, and extreme temperatures. Parasite DNA was extracted from DBS and used for PCR-based analysis to differentiate reinfection from recrudescence as well as sequencing of key mutations. The parasite DNA was extracted from three 3 mm punches from DBS samples using QIAamp DNA blood mid-kit (Qiagen GmbH, Hilden, Germany) according to the manufacturer’s instructions. Paired samples (day 0 and day of parasite recurrence) were genotyped using merozoite surface proteins 1 and 2 (*msp1* and *msp2*), and glutamate rich protein (*glurp*) genes as previously described, based on length polymorphisms on agarose gels. Gel analyser version 19.1 was used to estimate fragment sizes, and PCR artefacts were manually removed. The bins used to define a match were 10 bp for *msp1/msp2*, and 50 bp for *glurp*. The WHO decision algorithm was used to classify recrudescence and new infection.^23^ A recurrent parasite was classified as recrudescence if at least one allele was shared at all three loci on day 0 and the day of parasite recurrence while a new infection was defined as the absence of a shared allele at any of the three loci between day 0 and the day of parasite recurrence. Inconclusive results were reported as non-determined and excluded from the analysis of treatment outcome.

#### Molecular markers of antimalarial drug resistance

Day 0 and DNA extracts from recurrent infections were analysed for the presence of single nucleotide polymorphisms (SNPs) in the K13 gene (codons 430-720) linked to artemisinin resistance^24^ and *P. falciparum* multidrug resistance 1 (*Pfmdr1)* genes (codons 86, 184, 1034, 1042, and 1246) suspected to be associated with resistance/reduced susceptibility to 4-aminoquinolines using capillary sequencing (Sanger method) according to the protocols adopted from US. Centre for Disease Control and Prevention (CDC), Atlanta, USA.^19^ SNP calls in *Pfmdr1* and K13 genes were performed using Geneious® analysis software version 2022.2.2 (Biomatters, New Zealand; www.geneious.com) by mapping the sequence data on the 3D7 reference sequences.^19^

DNA samples of 49 K13 561H (mutant) and 34 K13 R561 (wildtype) parasites were used to conduct whole genome sequencing (WGS) to assess extended haplotypes around the gene. The DNA underwent selective whole genome amplification (sWGA) in three separate reactions as previously described.^25^ The sWGA products were combined into a single tube and underwent library preparation for Illumina sequencing using the Adapterama protocol. Prepared libraries were normalised, pooled and sequenced using 2X150bp chemistry on a NovaSeq6000 at the University of North Carolina High Throughput Sequencing Facility. We downloaded publicly available WGS data (n = 25) from *P. falciparum* isolates collected in 2014/15 in Rwanda^26^, Kagera region Tanzania (n = 9) ^21^, and Southeast Asian (SEA; n = 74) *P. falciparum* isolates carrying 561H mutants from Pf7K website (<u>Pf7: an open dataset of Plasmodium falciparum</u> <u>genome variation in 20,000 worldwide samples)</u>.^27^ The reads were processed using GATK4 following previously published methods (https://github.com/Karaniare/Optimized_GATK4_pipeline)^28^ and combined with data from Kagera, Tanzania in the 2021 study^21^, Rwanda and SEA and did extended haplotypes plot using the ggplot2 R package.

#### Treatment outcome

Treatment outcomes were classified as either early treatment failure (ETF), late clinic a l treatment failure (LCF), late parasitological treatment failure (LPF) or adequate clinical and parasitological response (ACPR) according to the WHO protocol.^2^ The primary endpoint of the study was PCR-adjusted ACPR at 28 days follow-up. Secondary endpoints included parasite clearance in terms of the presence of parasites at day 3 and parasite clearance time (PCT) 72 hours post-treatment, polymorphism in K13 and *Pfmdr1* genes, and the occurrence of adverse events.

#### Sample size estimation

The sample size was determined with the assumption that 5% of patients were likely to have treatment failure with either AL or ASAQ. With a confidence level of 95% and an estimated precision of 5%, the minimum sample size was 73 patients in each group. With an allowance of 20% to account for loss to follow-up and withdrawals during the 28-day follow-up, the target was 88 patients per drug.

#### Data management and analysis

Data were double-entered into a Microsoft Access database by two independent data clerks, validated, cleaned, and analysed using STATA for Windows, version 17 (STATA Corporation, TX-USA). Data was also transferred to the WHO Excel software programme (https://www.who.int/malaria/areas/treatment/drug_efficacy/en/) for management and analysis.^2^ Treatment outcomes were analysed using both per-protocol and Kaplan–Meier methods. Patients who were lost to follow-up or withdrawn during follow-up or who had reinfection or indeterminate PCR results were excluded from the PCR-corrected per-protocol analysis. For the Kaplan–Meier analysis, patients who were lost to follow-up or withdrawn during follow-up were censored on the last day of follow-up, patients with a new infection were censored on the day of reinfection, and patients with undetermined PCR results were excluded. Descriptive statistics, including proportion, mean, standard deviation, and range, were reported as appropriate. Chi-square tests were used to compare categorical data. Continuous variables such as parasite density and age were compared using the t-test (for normally distributed data) or the Mann-Whitney U-test (a non-parametric test for non-normally distributed data). Confidence intervals were calculated for binomial proportions. Two-sided p-values of less than 0.05 were considered statistically significant.

Analysis of parasite clearance was performed using the parasite clearance estimator as previously described^29^. The minimum detectable parasitaemia was set at 16 asexual parasites/ µl of blood and samples with too few data points and/or too low parasitaemia were excluded. The estimates which were generated included clearance rate constant (estimated clearance rate constant/hr), slope half-life (time in hrs needed for parasitaemia to decline by half, considering initial parasitaemia and excluding lag phase and the tail), and P50 and P95 (estimated time in hrs for the parasitaemia to decline by 50% and 95% of the initial values, respectively). The analysis was initially done for all patients and thereafter, it was repeated with only patients who had parasitaemia on day 3 and/or 561H mutations in K13 gene. All patients with slope half-life >5.0 hrs were considered to have delayed parasite clearance time as previously described.^29^

#### Ethical Considerations

Ethical clearance was obtained from the Medical Research Coordinating Committee of the National Institute for Medical Research (NIMR-MRCC) and WHO ethics review committee. Additional permission to conduct the study at Bukangara dispensary was obtained from the President’s Office, Regional Administration and Local Government (PO-RALG) as well as regional and district medical authorities. Parents/guardians of potential study children were informed about the objectives and procedures of the study and provided a written informed consent prior to recruitment. Study children obtained free health care throughout the follow-up period and travel expenses for follow-up at health facilities were covered.

## Results

Of the 343 patients screened, 176 (51.3%) were sequentially enrolled starting with AL (n=88) and then with ASAQ (n=88). The children in both treatment groups had comparable baseline characteristics (p>0.05), including age, anthropometric measurements, Hb level, axillary temperature and parasitaemia (Table 1).

**Table 1.**
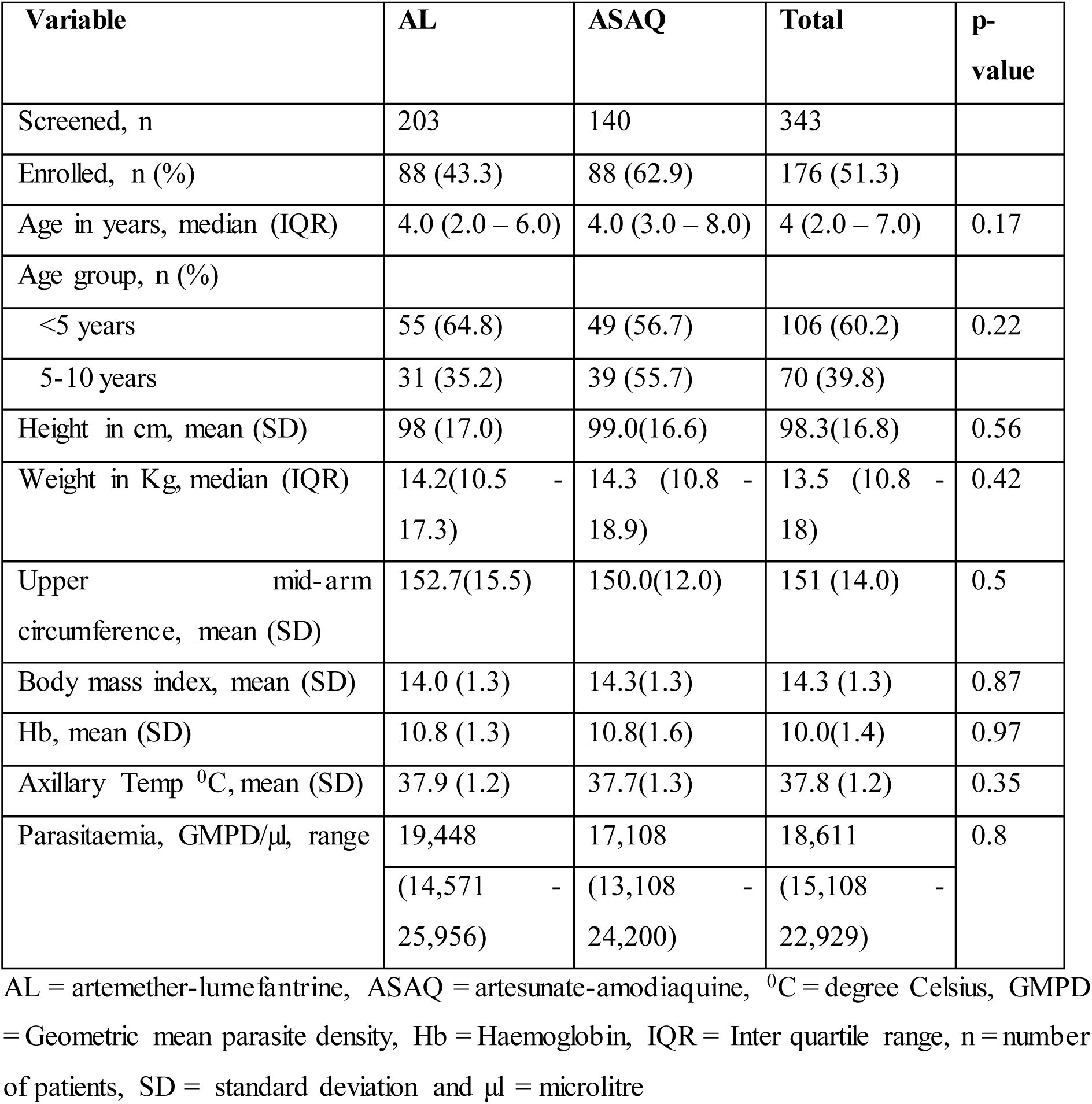
Baseline characteristics of the study patients.

| Variable | AL | ASAQ | Total | p-value |
| --- | --- | --- | --- | --- |
| Screened, n | 203 | 140 | 343 |  |
| Enrolled, n (%) | 88 (43.3) | 88 (62.9) | 176 (51.3) |  |
| Age in years, median (IQR) | 4.0 (2.0 – 6.0) | 4.0 (3.0 – 8.0) | 4 (2.0 – 7.0) | 0.17 |
| Age group, n (%) |  |  |  |  |
| <5 years | 55 (64.8) | 49 (56.7) | 106 (60.2) | 0.22 |
| 5-10 years | 31 (35.2) | 39 (55.7) | 70 (39.8) |  |
| Height in cm, mean (SD) | 98 (17.0) | 99.0(16.6) | 98.3(16.8) | 0.56 |
| Weight in Kg, median (IQR) | 14.2(10.5 - 17.3) | 14.3 (10.8 - 18.9) | 13.5 (10.8 - 18) | 0.42 |
| Upper mid-arm circumference, mean (SD) | 152.7(15.5) | 150.0(12.0) | 151 (14.0) | 0.5 |
| Body mass index, mean (SD) | 14.0 (1.3) | 14.3(1.3) | 14.3 (1.3) | 0.87 |
| Hb, mean (SD) | 10.8 (1.3) | 10.8(1.6) | 10.0(1.4) | 0.97 |
| Axillary Temp °C, mean (SD) | 37.9 (1.2) | 37.7(1.3) | 37.8 (1.2) | 0.35 |
| Parasitaemia, GMPD/µl, range | 19,448 | 17,108 | 18,611 | 0.8 |
|  | (14,571 - 25,956) | (13,108 - 24,200) | (15,108 - 22,929) |  |

AL = artemether-lumefantrine, ASAQ = artesunate-amodiaquine, °C = degree Celsius, GMPD = Geometric mean parasite density, Hb = Haemoglobin, IQR = Inter quartile range, n = number of patients, SD = standard deviation and µl = microlitre

All 176 study children completed follow-up with no loss to follow-up or withdrawal, and all had treatment outcomes assigned (Supplementary Fig 1). Prior to PCR correction, 5 (4.6%) and 26 (30.7%) of the AL-treated children had LCF and LPF, respectively (Table 2). These parasite recurrences occurred on day 21 (54.8%, n=17) and day 28 (35.5%, n=11) and the remaining three (9.7%) on day 14. In the ASAQ group, 1 (1.1%) LCF and 1 (1.1%) LPF were observed. The PCR-uncorrected ACPR was 64.8% (95% CI: 53.9 - 74.7%) and 97.8% (95% CI: 92.0 - 99.7%) in the AL and ASAQ groups, respectively. After PCR correction, the ACPR per protocol was 98.3% (95% CI: 90.8 - 100%) for AL and 100% (95% CI: 95.8 - 100%) for ASAQ (Table 2). Kaplan-Meier analysis showed a similar cure rate of 98.3% for AL and 100% for ASAQ.

**Table 2.** PCR uncorrected and corrected treatment outcomes.

| Treatment outcome | Artemether-lumefantrine |  | Artesunate-amodiaquine |  |
| --- | --- | --- | --- | --- |
|  | N(%) | CI 95% | N(%) | CI 95% |
| *PCR uncorrected* |  |  |  |  |
| ETF<sup>a</sup>, n(%) | 0 | 0 | 0 | 0.0-4.1 |
| LCF<sup>b</sup>, n(%) | 5(5.7) | 1.9-12.8 | 1(1.1) | 0.0-6.2 |
| LPF<sup>c</sup>, n(%) | 26(29.5) | 20.3-40.2 | 1(1.1) | 0.0-6.2 |
| ACPR<sup>d</sup>, n(%) | 57(64.8) | 53.9-74.7 | 86(97.7) | 92.0-99.7 |
| Total patients per protocol | 88 |  | 88 |  |
| KM: cumulative cure rate | 57(64.8) | 53.8-73.7 | 86(97.7) | 91.2-99.4) |
| *PCR corrected* |  |  |  |  |
| ETF<sup>a</sup>, n(%) | 0 | 0.0-6.2 | 0 | 0.0-4.2 |
| LCF<sup>b</sup>, n(%) | 0 | 0.0-6.2 | 0 | 0.0-4.2 |
| LPF<sup>c</sup>, n(%) | 1(1.87) | 0.0-9.2 | 0 | 0.0-4.2 |
| ACPR<sup>d</sup>, n(%) | 57(98.3) | 90.8-100 | 86(100) | 95.8-100 |
| Total patients per protocol | 58 |  | 86 |  |
| New infections | 28 |  | 2 |  |
| Non-determinant | 2 |  |  |  |
| KM: cumulative cure rate | 57(98.3) | 90.8-100 | 86(100) | n/a |

ETF<sup>a</sup> = early treatment failure, LCF<sup>b</sup> = late clinical failure, LPF<sup>c</sup> = late parasitological failure and ACPR<sup>d</sup> = adequate clinical and parasitological response

At day 3 (72 hours post-treatment), parasitaemia was detected by microscopy in 12.5% (11/88) of the patients in the AL group and in 19.3% (17/88) in the ASAQ-treated group. When analysing parasite clearance half-life, four patients (2.3%) were excluded (one in the AL and 3 in the ASAQ group) because they had too few data points, leaving 172 patients (97.7%). The assessment of parasite clearance time based on various estimates (clearance rate constant, slope half-life, P50 and P90) was similar for both drugs (Table 3).

**Table 3:** Parasite clearance time among patients treated with AL and ASAQ.

| **Variables (all patients)** | **AL (n=87)<sup>a</sup>** | **ASAQ (n=85)<sup>b</sup>** | **Total (N=172)** |
| --- | --- | --- | --- |
| **(all patients)** |  |  |  |
| Clearance rate constant (IQR)/(hrs) | 0.18(0.13,0.26) | 0.18(0.11,0.23) | 0.18(0.12,0.25) |
| Slope half-life (hrs) (IQR) | 3.77(2.67,5.51) | 3.93(2.97,6.03) | 3.88(2.77, 5.83) |
| P50 (hrs)<sup>c</sup> | 9.29(4.22,14.83) | 5.78(3.29,12.72) | 7.83(3.86,14.29) |
| P95 (hrs)<sup>d</sup> | 21.38(16.83,30.80) | 21.02(14.67,31.95) | 21.07(15.50,30.98) |
| **Patients with day 3 parasitaemia and *k-13* mutations** | **AL (n=24)** | **ASAQ (n=25)** | **Total (N=49)** |
| Clearance rate constant (IQR)/(hrs) | 0.11(0.10,0.12) | 0.09(0.08, 0.11) | 0.10(0.08, 0.12) |
| Slope half-life (hrs) (IQR) | 6.54(5.80,7.08) | 7.40(6.12,9.04) | 6.71(5.92, 8.16) |
| P50 (hrs)<sup>c</sup> | 14.03(9.13, 19.58) | 14.30(8.34, 19.85) | 14.30(8.79, 19.64) |
| P95 (hrs)<sup>d</sup> | 34.41(29.22,44.88) | 38.54(32.26,45.11) | 37.32(29.42,45.05) |

<sup>a</sup>One patient in AL and <sup>b</sup>three patients in ASAQ were excluded because they had too few data.
<sup>c</sup>Estimated time (hrs) for parasitaemia to decline by 50% of the initial values and <sup>d</sup>Estimated time (hrs) for parasitaemia to decline by 95% of the initial values

The commonly reported adverse events included cough, runny nose, abdominal pain, fever, nausea and diarrhoea (Table 4), and most of these events were reported in the ASAQ-treated group. There was no SAE and the AEs were mild and further analysis showed that they were not related to the study drugs.

**Table 4:** Adverse events reported among patients treated with AL and ASAQ.

| Type of adverse event | AL (n=88) | ASAQ (n=88) | Total (n=176) |
| --- | --- | --- | --- |
|  | n(%) | n(%) | n(%) |
| Cough | 29(32.9) | 29(32.9) | 58(32.9) |
| Runny nose | 3(3.4) | 13(14.8) | 16(9.1) |
| Abdominal pain | 2(2.3) | 11(12.5) | 13(7.4) |
| Fever | 3(3.4) | 10(11.4) | 13(7.4) |
| Nausea | 1(1.1) | 3(3.4) | 4(2.3) |
| Watery diarrhoea | 1(1.1) | 1(1.1) | 2(1.1) |
| Loose stool | 0(0) | 2(2.3) | 2(1.1) |
| Loss appetite | 1(1.1) | 1(1.1) | 2(1.1) |
| Sling pale | 0(0) | 1(1.1) | 1(0.6) |
| Urination | 0(0) | 1(1.1) | 1(0.6) |
| Vomiting | 1(1.1) | 0(0) | 1(0.6) |
| Bee sting on the face | 1(1.1) | 0(0) | 1(0.6) |
| **Total** | **42(47.7)** | **72(81.8)** | **114(64.8)** |

Table 5 shows the results of the molecular marker analysis for the K13 and *Pfmdr1* genes. The K13 gene was successfully sequenced in 87 (98.9%) and 86 (97.7%) of the AL and ASAQ pre-treatment samples (day 0), respectively. In addition, 29 (93.5%) and 2 (100%) of the AL and ASAQ post-treatment samples with recurrent parasitaemia, respectively, yielded interpretable results (Table 5). Overall, 22.5% (39/173) of day 0 samples had a validated 561H mutation associated with ART-R, and of these, 24.1% and 20.9% were in the AL and ASAQ groups, respectively. The majority of patients with persistent parasitaemia on day 3 had a 561H mutation, 72.7% (8/11) in the AL and 58.8% (10/17) in the ASAQ group. The 561H mutation was significantly associated with parasitaemia on day 3 in both treatment groups (p <0.001), but not with recurrent infections (in the AL group (p=0.790). In both treatment groups, the parasite clearance half-life was >6.5 hours in patients with parasitaemia on day 3 and/or 561H mutation, exceeding the threshold of >5 hrs seen in parasites with artemisinin resistance. For the region 1 of *Pfmdr1* gene, all samples were successfully analysed for both drugs. All samples carried the N86Y wild type, while the Y184F mutation was detected in 42% of day 0 in both treatment groups. For *Pfmdr1* mutations in region 2, 92% and 46.7% of samples in the AL and ASAQ groups, respectively, yielded interpretable results. Of these, four samples (three samples on day 0 and one sample on the day of parasite recurrence) in the AL treatment group had mutations at codon D1246Y (Table 5).

**Table 5:**
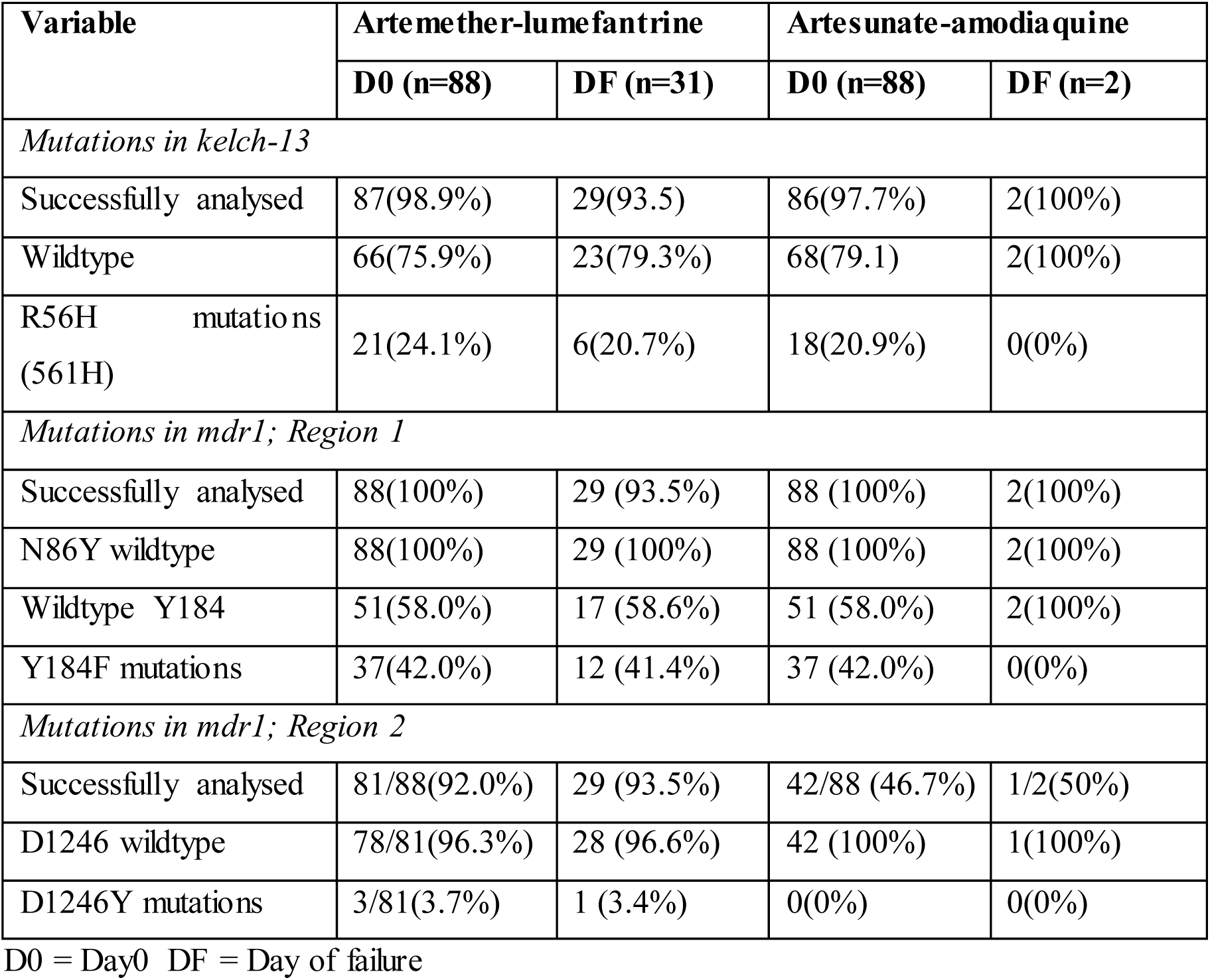
Mutations in *k13* and *Pfmdr1* genes.

Extended haplotype analysis was done to examine the genomes of this TES, MSMT 2021, Rwanda, and SEA *P. falciparum* isolates. The analysis showed that the TES 561H samples shared an extended haplotype flanking the mutation with the same pattern of variation within a core 9kb region around 561H as the previously described TZ2 haplotype from 2021 Kagera samples.^21^ Beyond the 9kb block, these TZ2 haplotypes diverge with multiple haplotype patterns consistent with recombination breaking down haplotypes quickly in this relatively high transmission region (Figure 2).

**Figure 2.**
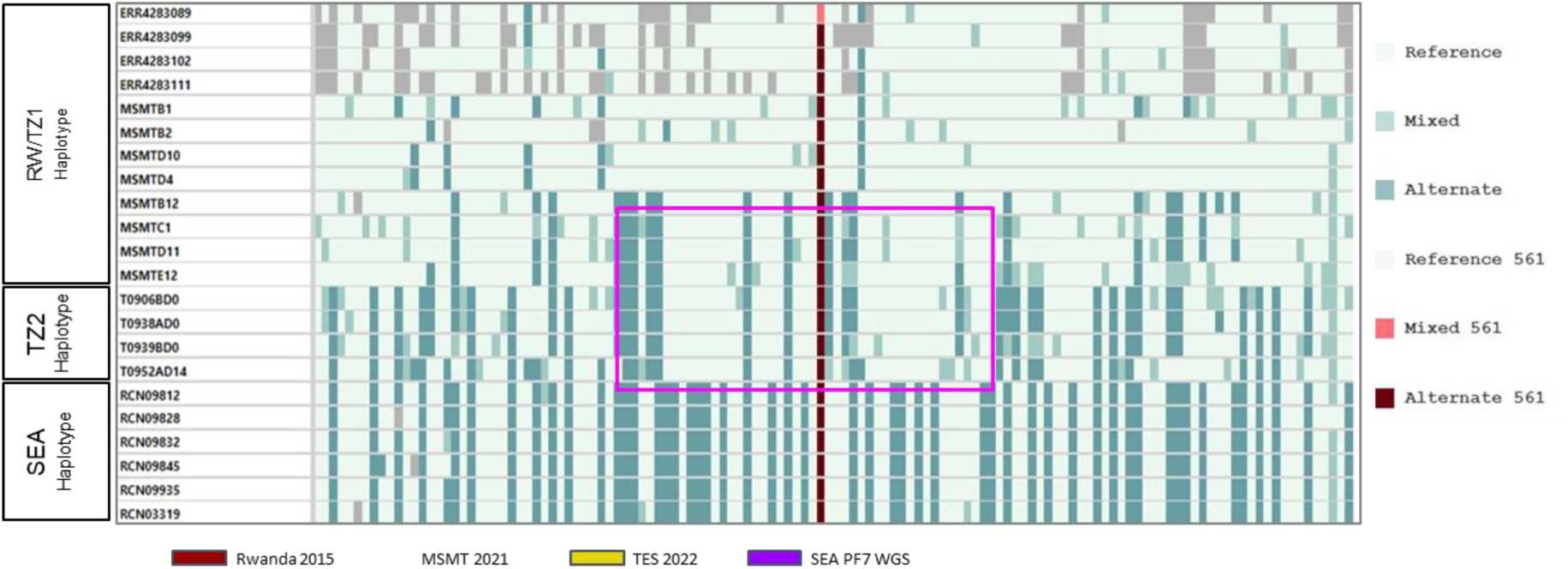
Extended flanking haplotype plot around K13 among single strain 561H mutants. The core haplotype described in Kagera in 2021 [haplotype two (TZ2)] is shown in the pink spanning approximately 9kb, while grey regions represent loci where a call could not be made. Sample nomenclature: TES 2022 (T09521D14, T0939BDO, T0938ADO, T0906BDO), 2021 Kagera (MSMTE12, MSMTD11, MSMTC1, MSMTB12, MSMTB1, MSMTB2, MSMTD10, MSMTD4), Rwanda 2014-2015 (ERR42833089, ERR42833099, ERR4283102, ERR4283111), and Southeast Asia (RC09812, RC09828, RC09832, RC09845, RC09935, RC03319).

## Discussion

The results of this study show that both AL and ASAQ are highly efficacious with PCR - corrected cure rates of 98.3% and 100%, respectively, supporting their continued use for the treatment of uncomplicated falciparum infection. However, delayed parasite clearance at day 3 was observed in 12.5% and 19.3% of the AL- and ASAQ-treated groups, respectively, above the 10% threshold for suspected artemisinin partial resistance.^3^ This delayed parasite clearance was significantly associated with the K13 561H mutation (≥58.8%) and the parasite clearance time of >6.5 hours, confirming for the first time the emergence of ART-R in Tanzania.

In addition to Tanzania, ART-R has also been confirmed in Rwanda with the R561H mutation, in Uganda with the A675V and C469Y mutations and in Eritrea with the R622I mutation.^12–14^ The emergence of ART-R in Africa is of great concern as more than 95% of malaria cases and deaths occur on the continent.^15^ The WHO recently launched a strategy to respond to antimalarial drug resistance in Africa and called on countries to develop a context-specific strategy to respond to this new threat.^9^ It is worth noting that the study site (Karagwe) borders both Rwanda and Uganda, where ART-R has recently been confirmed. The district is about 100 km from the Rwandan site (Rukara in Kayonza district), where ART-R with the 561H mutation was reported in 2015 and 2018.^12^ Cross-border collaboration between these countries (and others in the Great Lakes region of Africa) in developing harmonised strategies to respond to antimalarial drug resistance would strengthen the overall response to the threat of antimalarial drug resistance.

The Karagwe site was not one of the national surveillance sites for monitoring antimalar ia l drug efficacy and resistance. However, a nationwide MMS study conducted in 2021 found a prevalence of 22.8% of the 561H mutation in patients of all ages treated at Bukangara dispensary in Karagwe district.^21^ This prompted the conduct of this study at this site, which eventually confirmed emergence of ART-R and detected similar prevalence of the 561H mutation. This points to the potential role of MMS in monitoring molecular markers to help target where drug efficacy and resistance studies are done.

The parasites with mutations reported in this study do not resemble those from SEA or Rwanda but are similar to a new haplotype (TZ2) which was previously reported in MMS 2021 data in the site.^21^ The result from haplotype analysis suggesting that haplotypes reported in the TES samples was less likely imported from SEA rather it indicates new haplotype (TZ2) which was first detected in 2021^21^ is expanding in the region.

The high rate of reinfection in patients treated with AL may be related to the high transmis s ion in the area. Other studies have also reported a similarly high rate of reinfection among AL-treated groups in areas of high transmission in the country.^18–20^ Therefore, efforts should be directed towards intensifying malaria control intervention to decrease the risk of malaria infections in the community. Such efforts are also crucial and should be part of the ART-R response strategy.

The results of the study demonstrate that the recommended first-line and alternative ACTs remain highly efficacious despite the presence of ART-R, which is consistent with previous reports from Africa^12–14^ and Southeast Asia.^3^ This suggests that ART-R does not compromise the efficacy of the ACT tested if the partner drug remains effective. However, ART-R puts partner drugs at greater risk by exposing an increased number of parasites to the partner drug alone.^30^ Continued monitoring of the efficacy of recommended ACTs and molecular markers of artemisinin and partner drugs at sentinel sites and beyond is critical. Lessons learned from recent nationwide MMS for K13 mutations should be used to identify hotspots for K13 mutations and determine where to conduct therapeutic efficacy studies.

## Data Availability

Once the publication process is completed, the clinical, laboratory and parasite sequencing data which have been shared with WHO will be available upon request to the corresponding author. The data include de-identified patient clinical data, a data dictionary and sWGS data. Requests to conduct analyses that are outside the scope of this publication will be reviewed by the core study team (DSI, CIM, MW, CR, JB and JJJ) to determine if the proposed use of the data is scientifically and ethically appropriate and falls within the permits of the institutions that provided ethical approval for the study. Any requests to reanalyse the data presented in this paper will not need to be reviewed by the MRCC of NIMR. The data have been shared with WHO and will be uploaded on its database.

## Abbreviations

ACPR: Adequate clinical and parasitological response
ACT: Artemisinin based combination therapy
AE: Adverse event
AL: Artemether- lumefantrine
ASAQ: Artesunate-amodiaquine
DBS: Dried blood spots on filter papers
ETF: Early treatment failure
*glurp*: Glutamate rich protein gene
K13: Kelch propeller domain located on Chromosome 13 of *P. falciparum*
LCF: Late clinical failure
LPF: Late parasitological failure
MRCC: Medical Research Coordinating Committee
*msp1*: Merozoite surface protein 1 gene
*msp2*: Merozoite surface protein 2 gene
NIMR: National Institute for Medical Research
NMCP: National Malaria Control Programme
PCR: Polymerase chain reaction
*Pfmdr1*: *Plasmodium falciparum* multidrug resistance 1 gene
SAE: Serious adverse event
WHO: World Health Organization

## Contributors

DSI, CIM, IG, RJAN and MW conceived, designed, and developed the study protocol. DSI, CIM, RAM, MDS, FF and CB implemented the field activities and/or laboratory analysis of samples. DSI, CIM, RJAN and JK supervised the implementation of the study and data analysis with technical support from MW and CR. DSI, AF, JB and JJJ planned the sequencing of samples which was done under JJJ’s supervision and AF performed the bioinformatics analys is. DSI, CIM, AF, JB, JJJ, MW and CR developed the manuscript and all authors reviewed the draft and approved the final version of the manuscript.

## Acknowledgements

The team is indebted to the children and their parents/guardians for accepting to take part in the study and attending the follow-up visits. Thanks to NIMR staff (August Nyaki, Gerion Gaudin, Oswald Oscar, Ezekiel Malecela, Hatibu Athumani, Thomas Semdoe, Twalipo Mponzi, Dativa Pereus, Rule Budodo, Salome Simba, Mwanaidi Mtui and Christopher Masaka) and those from Bukangara dispensary (Wildon Rwakilomba, Agness Kyovecho, Alinala Mgoye, Genoveva Joel, Nasra Asimwe and Esres Kakuru) for taking part in the implementation of the study at different stages. The support provided by the regional and district authorities (particularly the Regional Administrative Secretary, Regional Medical Officer, Regional Malaria Focal Person, the District Commission, the District Executive Directors, District Medical Officer and the District Malaria Focal Person) is highly appreciated. We are grateful to NIMR Management and officials from NMCP and PO-RALG for supporting the project. We thank Dr. Pascal Ringwald for his technical support. Permission to publish the manuscript was sought and obtained from the Director General of NIMR.

This work was supported, in part, by the Bill & Melinda Gates Foundation through WHO (OPP1209843) and NIMR [grant number 002202]. Under the grant conditions of the Foundation, a Creative Commons Attribution 4.0 Generic License has already been assigned to the Author Accepted Manuscript version that might arise from this submission. Funding was also received from US National Institute for Health (R01AI156267 to JAB, DSI and JJJ and K24AI134990 to JJJ).

