## Supplementary Figure 1, Supplementary Table 1 and Supplementary Table 2 for "Evidence of artemisinin partial resistance in North-western Tanzania: clinical and drug resistance markers study"

**Supplementary Figure 1: Trial profile showing the flow of patients during screening, enrolment and follow-up**

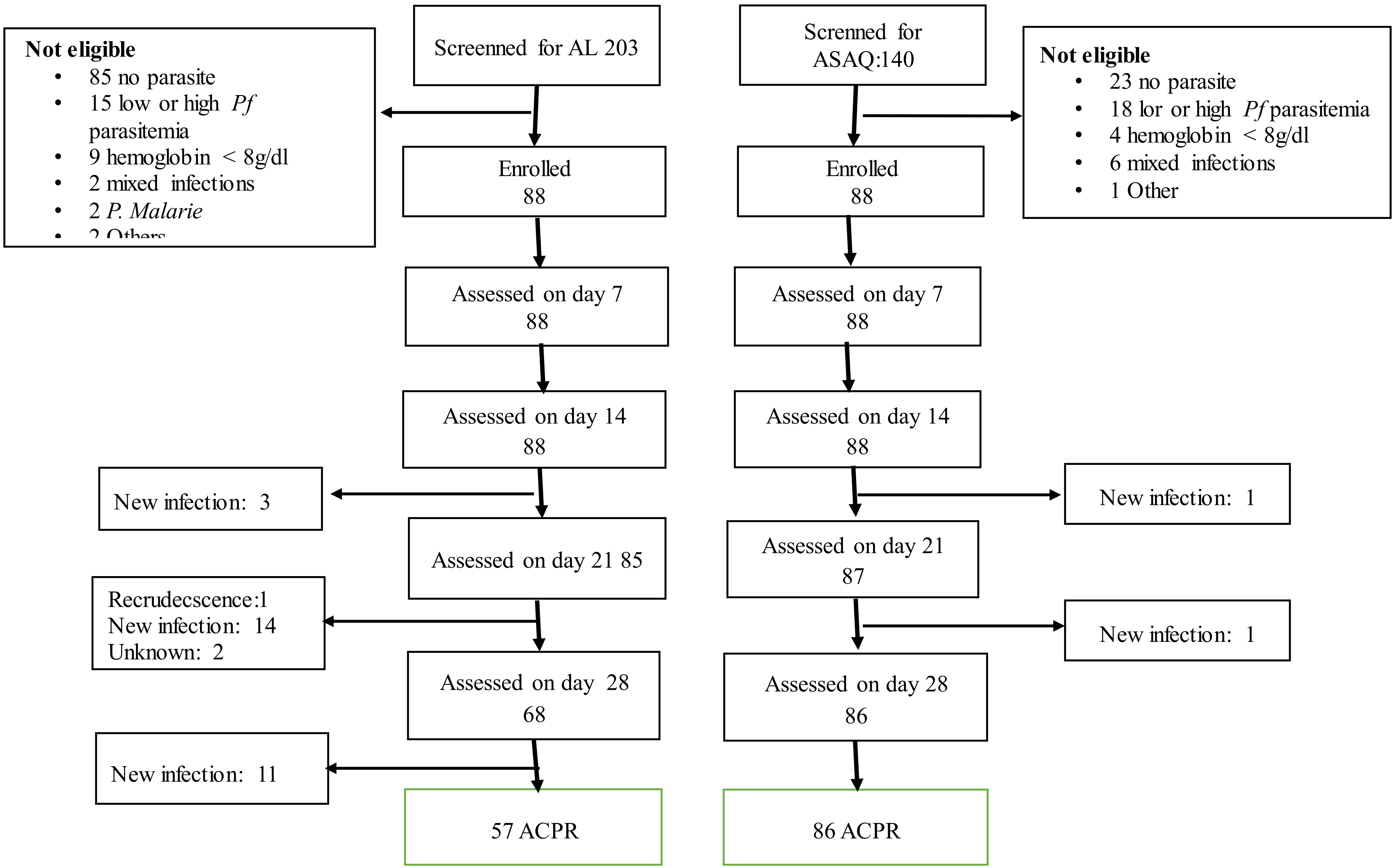

**Supplemental Table 1: AL_genotyping results**

| ***SAMPLE ID*** | ***MSP1*** | | | | ***MSP2*** | | | ***GLURP*** | | |
|---|---|---|---|---|---|---|---|---|---|---|
| | ***K1*** | ***Mad20*** | ***RO33*** | ***Result; msp1*** | ***IC1*** | ***FC27*** | ***Result; msp2*** | | ***Result;glurp*** | ***Result;all markers*** |
| T0905A D0 | 254 | 200 | 0 | R | 457 | 487 | NI | 929 | NI | NI |
| T0905A D28 | 246 | 0 | 0 | | 369 | 0 | | 1023 | | |
| T0906A D0 | 225 | 0 | 0 | R | 498 | 638 | R | 1177 | R | R |
| T0906A D21 | 220 | 0 | 113 | | 502 | 0 | | 1146 | | |
| T0908A D0 | 217 | 232 | 120 | R | 453 | 656 | NI | 994 | NI | NI |
| T0908A D28 | 0 | 0 | 122 | | 561 | 0 | | 1175 | | |
| T0912A D0 | 231 | 236 | 0 | NI | 480 | 625 | NI | 1302 | NI | NI |
| T0912A D21 | 257 | 0 | 0 | | 441 | 0 | | 1215 | | |
| T0914A D0 | 244 | 0 | 116 | NI | 473 | 673 | NI | 1181 | NI | NI |
| T0914A D21 | 0 | 194 | 0 | | 629 | 0 | | 1285 | | |
| T0915A D0 | 271 | 0 | 126 | NI | 449 | 557 | NI | 1181 | R | NI |
| T0915A D21 | 205 | 0 | 0 | | 0 | 646 | | 1194 | | |
| T0918A D0 | 258 | 0 | 128 | NI | 0 | 714 | NI | 1190 | NI | NI |
| T0918A D21 | 239 | 0 | 0 | | 0 | 643 | | 1371 | | |
| T0924A D0 | 263 | 228 | 0 | R | 515 | 624 | NI | 986 | NI | NI |
| T0924A D14 | 265 | 0 | 0 | | 0 | 674 | | 1445 | | |
| T0925A D0 | 0 | 0 | 137 | NI | 511 | 0 | NI | 1213 | R | NI |
| T0925A D21 | 260 | 0 | 0 | | 484 | 686 | | 1229 | | |
| T0926A D0 | 283 | 0 | 0 | NI | 460 | 684 | NI | 1200 | NI | NI |
| T0926A D28 | 0 | 241 | 0 | | 535 | 0 | | 1272 | | |
| T0933A D0 | 240 | 0 | 152 | NI | 499 | 608 | NI | 1230 | NI | NI |
| T0933A D28 | 271 | 303 | 0 | | 723,605 | 591 | | 1160 | | |

| | | | | | | | | | | |
|---|---|---|---|---|---|---|---|---|---|---|
| T0934A D0 | 311 | 294 | 0 | | 0 | 713 | | 1228 | | |
| T0934A D28 | 322 | 0 | 0 | NI | 496 | 0 | NI | 1169 | NI | NI |
| T0935A D0 | 325 | 0 | 0 | | 522 | 602 | | 829 | | |
| T0935A D28 | 0 | 0 | 165 | NI | 0 | 560 | NI | 1028 | NI | NI |
| T0936A D0 | 0 | 245 | 166 | | 557 | 624 | | 1355 | | |
| T0936A D28 | 370 | 0 | 166 | R | 567 | 633 | R | 1147 | NI | NI |
| T0937A D0 | 247 | 272 | 0 | | 584 | 0 | | 1177 | | |
| T0937A D28 | 0 | 0 | 173 | NI | 634 | 611 | NI | 1091 | NI | NI |
| T0939A D0 | 0 | 261 | 0 | | 595 | 0 | | 1373 | | |
| T0939A D28 | 0 | 0 | 164 | NI | 652,558 | 0 | NI | 1131 | NI | NI |
| T0944A D0 | 202 | 0 | 0 | | 0 | 616 | | 1009 | | |
| T0944A D21 | 251 | 264 | 0 | NI | 431 | 541 | NI | 1057 | R | NI |
| T0949A D0 | 228 | 0 | 0 | | 0 | 548 | | 1204 | | |
| T0949A D21 | 222 | 244 | 0 | R | 0 | 526 | NI | 1230 | R | NI |
| T0950A D0 | 0 | 254 | 0 | | 0 | 454 | | 940 | | |
| T0950A D21 | 234 | 0 | 0 | NI | 433 | 0 | NI | 1033 | NI | NI |
| T0951A D0 | 0 | 213 | 0 | | 514 | 415 | R | 1042 | NI | NI |
| T0951A D28 | 0 | 0 | 131 | NI | 573,510 | 0 | NI | 898 | NI | NI |
| T0952A D0 | 214 | 274 | 301 | | 630 | 566 | | 917 | | |
| T0952A D14 | 208 | 0 | 0 | R | 0 | 530 | NI | 1037 | NI | NI |
| T0953A D0 | 234 | 303 | 123 | | 489 | 0 | | 1035 | | |
| T0953A D28 | 225 | 296 | 302 | R | 666 | 537 | NI | 948 | NI | NI |
| T0958A D0 | 283 | 0 | 131 | | 0 | 547 | | 1052 | | |
| T0958A D28 | 209 | 0 | 0 | NI | 632 | 0 | NI | 1077 | R | NI |
| T0964A D0 | 237 | 219 | 0 | | 609 | 454 | | 987 | | |
| T0964A D21 | 0 | 225 | 0 | R | 661 | 0 | NI | 0 | ND | NI |
| T0966A D0 | 262 | 0 | 0 | | 670 | 0 | | 1003 | | |
| T0966A D21 | 238 | 0 | 151 | NI | 611 | 0 | NI | 1005 | R | NI |
| T0967A D0 | 245 | 299 | 0 | ND | 518,485 | 582 | ND | 990 | ND | ND |

| | | | | | | | | | | |
|---|---|---|---|---|---|---|---|---|---|---|
| T0967A D21 | 0 | 0 | 0 | | 0 | 0 | | 0 | | |
| T0968A D0 | 0 | 366 | 171 | | 0 | 498 | | 1017 | | |
| T0968A D14 | 0 | 0 | 159 | NI | 0 | 491 | R | 964 | R | NI |
| T0977A D0 | 303 | 0 | 0 | | 648 | 589 | | 1053 | | |
| T0977A D21 | 0 | 0 | 169 | NI | 531 | 0 | NI | 1020 | R | NI |
| T0981A D0 | 252 | 0 | 0 | | 658 | 534 | | 1018 | | |
| T0981A D21 | 0 | 0 | 0 | ND | 0 | 0 | ND | 0 | ND | ND |
| T0982A D0 | 244 | 291 | 0 | | 0 | 604 | | 992 | | |
| T0982A D21 | 0 | 285 | 366 | R | 605 | 0 | NI | 965 | NI | NI |
| T0984A D0 | 263 | 0 | 0 | | 718 | 627 | | 922 | | |
| T0984A D21 | 290 | 0 | 0 | NI | 643 | 606 | NI | 860 | NI | NI |

**Supplemental Table 2: ASAQ_genotyping results**

| ***SAMPLE ID*** | ***MSP1*** | | | | ***MSP2*** | | | ***GLURP*** | | |
|---|---|---|---|---|---|---|---|---|---|---|
| | ***K1*** | ***Mad20*** | ***RO33*** | ***Result; msp1*** | ***IC1*** | ***FC27*** | ***Result; msp2*** | | ***Result;glurp*** | ***Result;all markers*** |
| T0928B D0 | 373 | 0 | 111 | | 405 | 437 | | 868 | | |
| T0928B D14 | 218 | 0 | 0 | NI | 470 | 0 | NI | 641 | NI | NI |
| T0978B_D0 | 292 | 0 | 0 | | 399 | 301 | | 941 | | |
| T0978B_D21 | 200 | 0 | 0 | ND | 619/565 | 0 | NI | 763 | NI | NI |
